# Amplitude-integrated EEG amplitudes differ between wakefulness and sleep states in children

**DOI:** 10.1101/2021.07.29.21261253

**Authors:** Verena T. Löffelhardt, Adela Della Marina, Sandra Greve, Hanna Müller, Ursula Felderhoff-Müser, Christian Dohna-Schwake, Nora Bruns

**Affiliations:** Department of Pediatrics I, Neonatology, Pediatric Intensive Care Medicine, and Pediatric Neurology, University Hospital Essen, University of Duisburg-Essen, Essen, Germany; TNBS, Centre for Translational Neuro- and Behavioral Sciences, University Hospital Essen, University of Duisburg-Essen, Essen, Germany; Department of Pediatrics, Neonatology and Pediatric Intensive Care, University of Marburg, Marburg, Germany

**Keywords:** amplitude-integrated EEG, sleep states, children, sleep, antiepileptic drugs, wakefulness

## Abstract

**Introduction:** Interpretation of pediatric amplitude-integrated EEG (aEEG) is hindered by the lack of knowledge on physiological background patterns in children. The aim of this study was to assess the amplitudes and bandwidths of background patterns during wakefulness and sleep in children from long-term EEGs.

**Methods:** Forty long-term EEGs from patients < 18 years of age without or only solitary interictal epileptiform discharges were converted into aEEGs. Upper and lower amplitudes (µV) of the C3 – C4, P3 – P4, C3 – P3, C4 – P4, and Fp1 – Fp2 channels were measured during wakefulness and sleep. Bandwidths (BW, µV) were calculated, and sleep states assessed during the episodes of interest. A sensitivity analysis excluded patients who received antiepileptic drugs.

**Results:** Median age was 9.9 years (interquartile range 6.1 – 14.7). All patients displayed continuous background patterns. Amplitudes and BW differed between wakefulness (C3-C4 channel: upper 35 (27 – 49), lower 13 (10 – 19), BW 29 (21 – 34)) and sleep. During sleep, episodes with high amplitudes (upper 99 (71 – 125), lower 35 (25 – 44), BW 63 (44 – 81)) corresponded to sleep states N2 – N4. These episodes were interrupted by low amplitudes that were the dominating background pattern towards the morning (upper 39 (30 – 51), lower 16 (11 – 20), BW 23 (19 – 31), sleep states REM, N1, and N2). With increasing age, amplitudes and bandwidths declined. The sensitivity analysis yielded no differences in amplitude values or bandwidths.

**Conclusion:** aEEG amplitudes and bandwidths were low during wakefulness and light sleep and high during deep sleep in stable children undergoing 24 hour EEG recordings. aEEG values were not altered by antiepileptic drugs in this study.

## Introduction

The use of amplitude-integrated electroencephalography (aEEG) to continuously monitor cerebral function is increasing in pediatric intensive care (1,2). Growing evidence in favor of continuous electroencephalography (cEEG) monitoring, limited access to full channel cEEG, and broad availability of aEEG devices in neonatal intensive care units promote this development (2-5). While aEEG monitoring is well-established in preterm infants and neonates, recent evidence suggests that aEEG helps to identify seizures in older children on the pediatric intensive care unit (PICU) (6-10). Furthermore, analysis of the background pattern may aid in predicting outcomes pediatric after cardiac arrest (11).

For interpretation of aEEG background patterns, the lack of reference values for infants and children constitutes a serious challenge for clinicians (1,2). A continuous background pattern with lower border values above 5-7 µV is generally considered normal (6,8,12). However, these values derive from a classification proposed for term born and preterm infants by Hellström-Westas which remains unvalidated in older infants or children (13).

Studies on normal values in newborns showed that variations of the upper and lower amplitudes concur with changes in wakefulness (13-16). These variations are referred to as sleep-wake cycling. The presence or absence of sleep-wake cycling is associated with outcomes in several diseases in neonates and young infants up to three months of age (17-19). In older infants and children, sleep patterns have been studied by 24-hour EEG and polysomnography. Amplitude height is used among other information to determine the sleep state from raw EEG, with lower amplitudes during light sleep or rapid-eye-movement (REM) sleep and higher amplitudes during deep sleep. Whether these amplitude variations in the raw EEG translate into a cycling pattern that can be recognized by aEEG has not yet been investigated.

Owed to the lack of evidence on physiological ranges and variations of amplitudes and bandwidth, aEEG interpretation in the pediatric intensive care setting mainly focuses on the detection of seizures and severe abnormalities (2,6-8,10). Guidance on the range of upper and lower amplitudes and bandwidth that can be expected during wakefulness and sleep in children could facilitate aEEG interpretation and promote the identification of abnormal electrocortical activity and sleep patterns. This retrospective single-centre study aimed to determine the ranges of upper and lower amplitudes in children during wakefulness and sleep from unremarkable long-term EEGs, which were converted into aEEGs prior to analysis.

## Methods

Long-term EEGs recorded between 2017 and 2020 in patients < 18 years of age in the Children’s Hospital of the University Hospital Essen without pathology or only solitary interictal epileptiform discharges (IEDs) were eligible for analysis. Pathological recordings with repeated IEDs or impaired sleep architecture were not considered for analysis. Patient histories were reviewed to extract clinical information.

### Ethics approval

The study was approved by the local ethics committee of the Medical Faculty of the University of Duisburg-Essen (20-9444-BO). Informed consent was not necessary according to local legislation, because data were anonymized.

### EEG recording

Upon admission, full channel EEG was applied according to the international 10-20 system after skin preparation with OneStep EEG Gel Abrasiv plus® and application of Elefix conductive EEG paste (Nihon Kohden). Impedance check was performed, and skin preparation repeated until impedances below < 50 kΩ were achieved for all electrodes. The parents or patients themselves recorded all activities during the recording. When in bed, additional video monitoring was performed until the next morning.

### aEEG analysis

The channels C3 – C4, P3 – P4, C3 – P3, C4 – P4, and Fp1 – Fp2 of the 10 – 20 system were converted into aEEGs using Polaris Trend Software QP-160AK (Nihon Kohden, Tokyo, Japan). VL and NB measured the upper and lower borders of amplitude of all channels semi-manually with the integrated tool for amplitude measurement.

During wakefulness, one hour of artifact-free recording in the afternoon was measured. For measurement of sleep amplitudes, the period between going to bed and five a.m. was considered. At that time, noises begin to rise in our hospital and sleep is disturbed. During sleep, amplitudes of the 30-minute section with the highest aEEG band were measured. For low amplitude measurement, the section with the lowest amplitudes during sleep was assessed. The measured sections were labelled and saved as a PDF file to enable identification of the exact same time interval for raw EEG review.

### Raw EEG review

Raw EEG review was performed by a board-certified pediatric neurologist (ADM) blinded to aEEG findings (certificates in EEG and epileptology by the German Society for Epileptology).

The time intervals identified from aEEGs during sleep were reviewed and the sleep state was determined according to the scoring system by the American Academy of Sleep Medicine (20). In case the sleep state changed during the reviewed interval, both sleep states were documented. As our recordings did not contain electro-oculography, REM sleep and N1 could not always be clearly differentiated. This was documented as “N1 or REM”.

When interictal epileptiform discharges were detected during the interval of interest, the recording was retrospectively excluded.

### Data analysis

Continuous variables are presented as median and interquartile range (IQR). The mean ± standard deviation (SD) is given where information on scattering of the values is of interest. The mean and 95% confidence intervals (CI) are given in the sensitivity analysis (see below) to enable comparison between groups without statistical hypothesis testing. 95% prediction limits (the range in which 95% of future measurements would be expected) were calculated based on the mean and standard deviation. For variables with non-normally distributed data, logarithmic transformation was performed before calculation of confidence intervals and prediction limits (upper and lower amplitudes and bandwidths of Fp1 – Fp2).

For discrete variables, absolute and relative frequencies are given.

#### Calculation of amplitudes and bandwidth

For each aEEG channel, we calculated the mean of the two measurements of the upper and lower amplitudes. The bandwidth was calculated as the difference between the upper and lower amplitudes.

#### Sensitivity analysis

A sensitivity analysis including only patients without antiepileptic drugs (AEDs) was performed for the upper and lower amplitudes and bandwidths.

#### Verification of measurements and interrater reliability

Systematic measurement differences between the two observers were ruled out using Bland-Altman plots (21). Outliers in the plots were assessed for typing errors, which were corrected if applicable. Next, a second set of Bland-Altman plots was created. This time, both observers reviewed the tracings of all outliers and found the measurements to be plausible in all cases. No changes were made to the original measurements.

To assess interrater reliability, we calculated intraclass correlation coefficients for the upper and lower amplitudes using a two-way mixed model for individual ratings (22).

#### Software

SAS Enterprise Guide 8.4 (SAS Institute Inc., Cary, NC, USA) was used to perform statistical analyses and produce figures.

## Results

Out of 98 performed long-term EEGs, 47 met the inclusion criteria. Seven cEEGs were excluded after raw EEG review due to abnormalities. The duration of the remaining 40 recordings ranged between 14:48 and 23:30 hours:minutes (median 20:11). Median age was 9.9 years (IQR 6.1 – 14.7 years) with 13 (33 %) male patients.

All patients were admitted to hospital electively in stable condition for the conduction of long-term cEEG. Twenty-two (55 %) cEEGs were conducted in patients with suspected seizures, 16 (40 %) in confirmed seizures or epilepsy, and two (5 %) after discontinuation of antiepileptic treatment. 16 (40 %) of patients received AEDs at the time of recording. Further information on patients, cEEG findings, and AEDs, is presented in table 1.

**Table 1:** Clinical information

|  | N (%)* |
| --- | --- |
| Age (years) [median (interquartile range)<br>mean ± standard deviation] | 9.9 (6.1 – 14.7)<br>9.9 ± 5.1 |
| Sex male | 13 (33 %) |
| Indications |  |
| Suspected seizures or epilepsy | 22 (55.0 %) |
| Treatment control | 16 (40.0 %) |
| Discontinuation of antiepileptic drugs | 2 (5.0 %) |
| Findings |  |
| Unremarkable | 26 (65.0 %) |
| Solitary focal IEDs | 6 (15.0 %) |
| Solitary generalized IEDs | 7 (17.5 %) |
| Solitary beta waves | 1 (2.5 %) |
| Antiepileptic drugs | 16 (40.0 %) |
| Monotherapy | 10 (25.0 %) |
| Combination therapy (2 drugs) | 5 (12.5 %) |
| Combination therapy (3 drugs) | 1 (2.5 %) |

\*except for age, IED = interictal epileptiform discharge

### aEEG patterns and sleep states

All tracings displayed continuous background patterns according to Hellström-Westas (13). We observed changes of the upper and lower amplitudes and bandwidth during the recording in all patients and in all analyzed channels (Table 2). The highest amplitudes and bandwidths occurred in the middle of the night, interrupted by short amplitude dips. Towards the morning, the amplitudes and bandwidths decreased (Figure 1).

**Figure 1:**
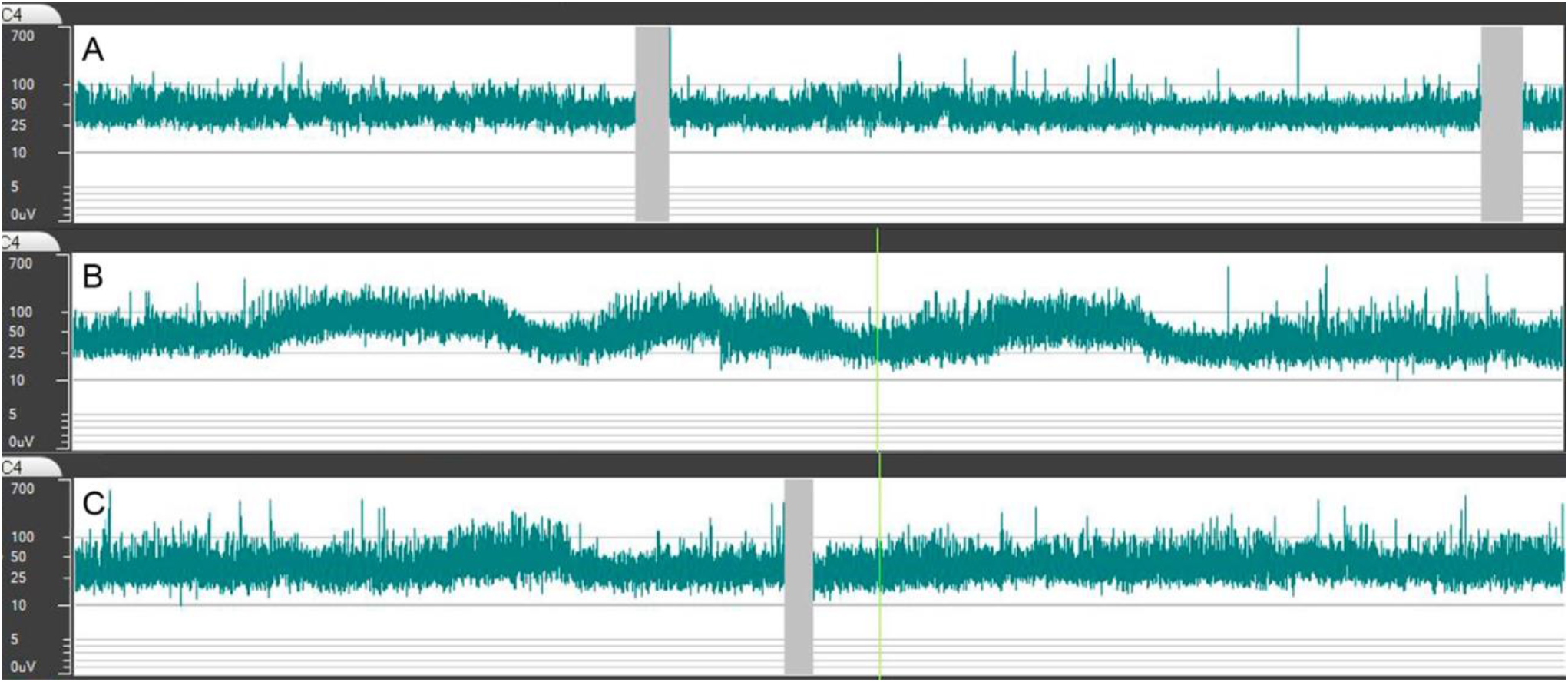
Typical course of amplitudes and bandwidth during recordings. A) aEEG during wakefulness with constant amplitudes and bandwidth. B) In the first half of the night, a rise in the upper and lower amplitude occurs. The bandwidth also increases but becomes less obvious due to the logarithmic scale of the aEEG. C) Towards the morning, the amplitudes and bandwidth lower again.

**Table 2:** Observed amplitude values and prediction limits for each channel

| Channel |  | Wakefulness |  |  | Sleep |  |  |  |  |  |
| --- | --- | --- | --- | --- | --- | --- | --- | --- | --- | --- |
| | | Lower ($\mu\text{V}$) | Upper ($\mu\text{V}$) | Bandwidth ($\mu\text{V}$) | High amplitude Lower ($\mu\text{V}$) | Upper ($\mu\text{V}$) | Bandwidth ($\mu\text{V}$) | Low amplitude Lower ($\mu\text{V}$) | Upper ($\mu\text{V}$) | Bandwidth ($\mu\text{V}$) |
| C3 – C4 | Median (IQR) | 13 (10 – 19) | 35 (27 – 49) | 29 (21 – 34) | 35 (25 – 44) | 99 (71 – 125) | 63 (44 – 81) | 16 (11 – 20) | 39 (30 – 51) | 23 (19 – 31) |
|  | 95% PL | 7 – 31 | 19 – 75 | 11 – 45 | 8 – 65 | 22 – 184 | 13 – 120 | 6 – 27 | 13 – 69 | 7 – 43 |
| P3 – P4 | Median (IQR) | 20 (16 – 25) | 49 (40 – 62) | 29 (23 – 38) | 34 (24 – 47) | 95 (68 – 124) | 59 (44 – 76) | 17 (13 – 20) | 42 (35 – 51) | 25 (22 – 31) |
|  | 95% PL | 9 – 31 | 23 – 79 | 13 – 48 | 9 – 63 | 26 – 168 | 16 – 106 | 8 – 26 | 21 – 67 | 12 – 42 |
| C3 – P3 | Median (IQR) | 16 (10 – 20) | 36 (26 – 46) | 23 (16 – 27) | 28 (18 – 36) | 82 (51 – 102) | 52 (33 – 62) | 12 (9 – 16) | 34 (25 – 44) | 21 (15 – 28) |
|  | 95% PL | 4 – 26 | 11 – 62 | 11 – 45 | 5 – 51 | 15 – 141 | 9 – 91 | 5 – 20 | 11 – 58 | 5 – 38 |
| C4 – P4 | Median (IQR) | 16 (11 – 21) | 39 (29 – 53) | 25 (17 – 30) | 28 (21 – 37) | 80 (56 – 104) | 52 (35 – 66) | 13 (10 – 16) | 36 (28 – 45) | 23 (17 – 28) |
|  | 95% PL | 5 – 28 | 13 – 67 | 8 – 40 | 4 – 55 | 11 – 154 | 7 – 100 | 5 – 21 | 13 – 60 | 7 – 40 |
| Fp1 - Fp2 | Median (IQR) | 18 (13 – 22) | 81 (57 – 93) | 59 (44 – 74) | 28 (21 – 34) | 85 (66 – 102) | 56 (45 – 68) | 10 (8 – 13) | 28 (22 – 34) | 17 (13 – 20) |
|  | 95% PL | 9 – 32 | 36 – 160 | 25 – 136 | 14 – 54 | 43 – 156 | 28 – 105 | 5 – 22 | 12 – 65 | 7 – 44 |

PL = prediction limits

High amplitudes were associated with sleep states N2 and N3, low amplitudes with wakefulness and sleep states REM, N1, and N2 (Figure 2). One patient was awake during the low amplitude episode at night. Amplitudes were slightly higher during wakefulness than during light sleep (table 2 and figure 3). With increasing age, amplitudes and bandwidths tended to be lower, especially during wakefulness and light sleep (Figure 4).

**Figure 2:**
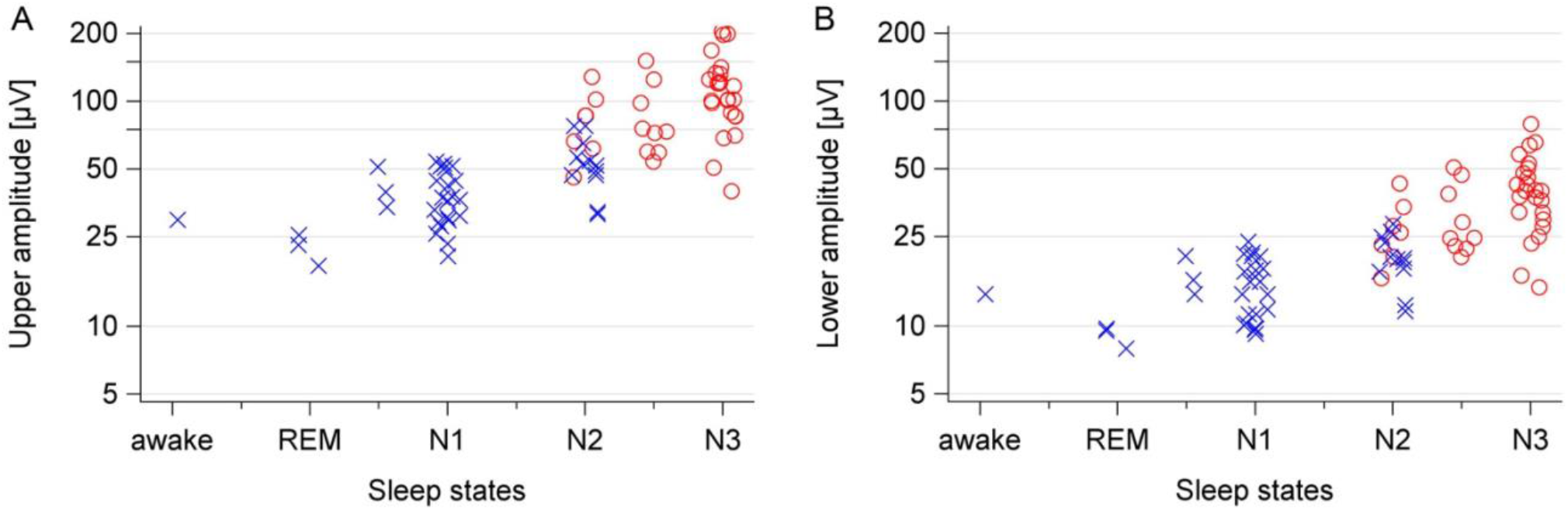
Distribution of diagnosed sleep states and the corresponding amplitudes. The colored symbols indicate if amplitudes were classified as high (red circle) or low (blue x) within the intraindividual tracing. The minor tick mark between N2 and N3 represents tracings with transition from N2 to N3 during the assessed time interval. Tracings that could not be clearly assigned to N1 or REM are plotted at the minor tick mark between the two states. A) Upper amplitude. B) Lower amplitude.

**Figure 3:**
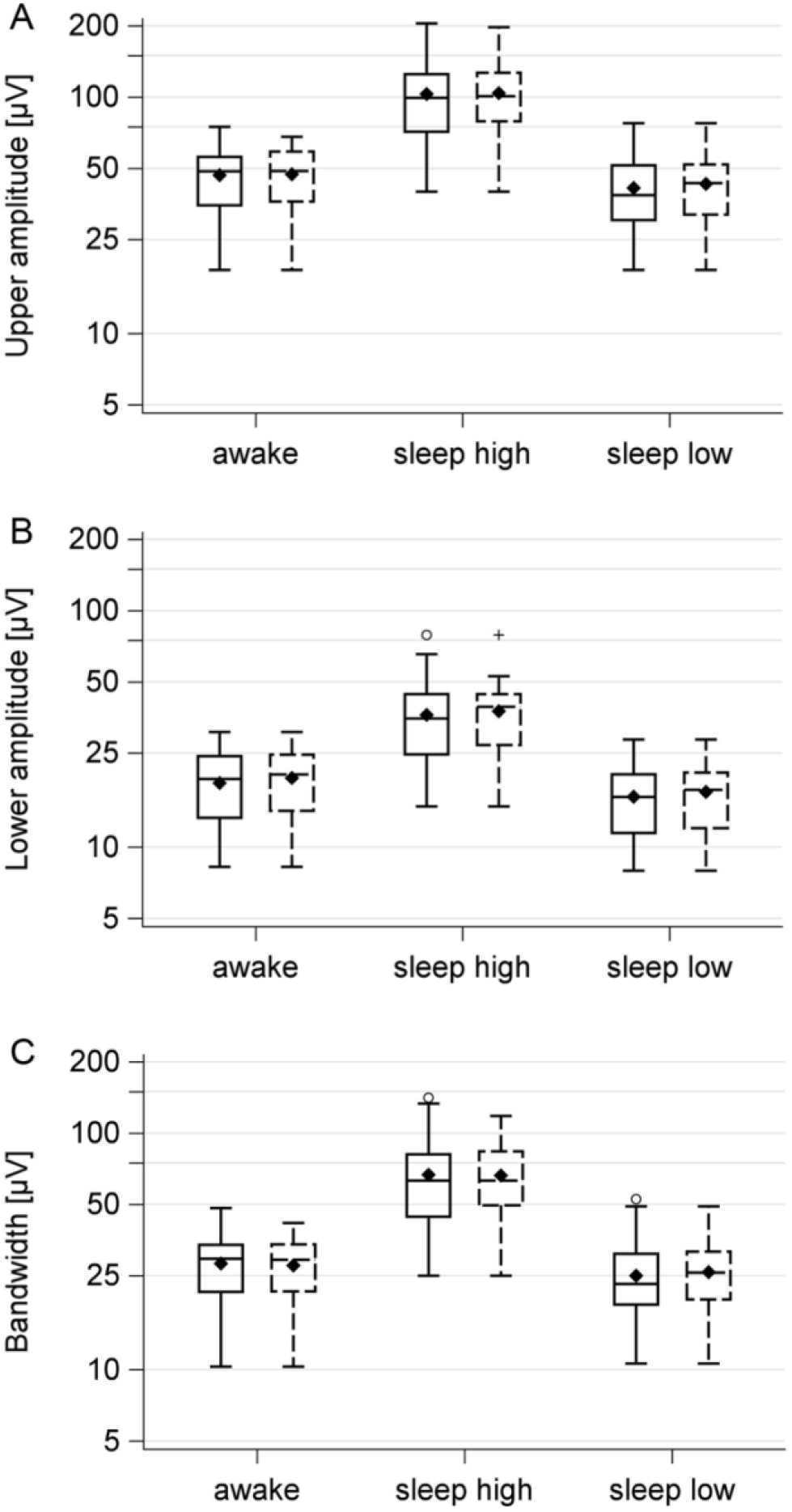
Sensitivity analysis of amplitudes and bandwidth. The solid boxes represent all patients, the dashed boxes patients without antiepileptic drugs. Whiskers show the 1.5 fold interquartile ranges, the mean is represented by the filled diamond. A) Upper amplitude. B) Lower amplitude. C) Bandwidth.

**Figure 4:**
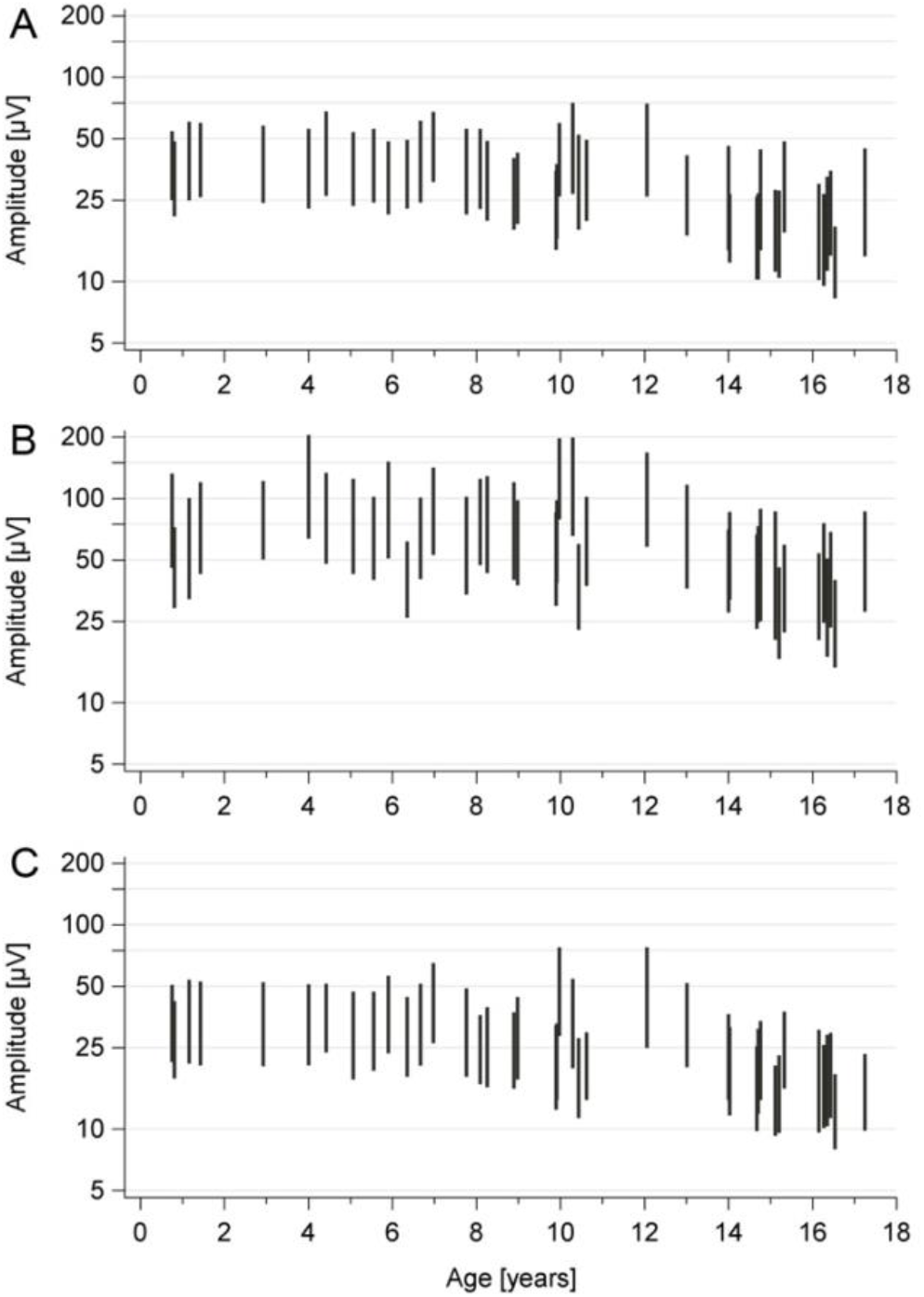
Upper and lower amplitudes and bandwidths of each individual tracing by age. A) During wakefulness. B) During the high amplitude sleep episode. C) During the low amplitude sleep episode.

The distribution of sleep states during low amplitude episodes did not differ between patients with and without AEDs. During high-amplitude sleep episodes, patients without AEDs displayed sleep state N2 in 2 (8.3 %), N2-N3 in 5 (20.8 %), and N3 in 17 (70.8 %) of cases. Patients receiving AEDs displayed N2 in 5 (31.3 %), N2-N3 in 4 (25 %), and N3 in 7 (43.8 %) of cases. Note that these numbers reflect the frequency of each sleep state during the assessed intervals, not the duration.

### Sensitivity analysis

The sensitivity analysis showed no differences in amplitude values and bandwidths between the whole study population and patients without AEDs (Figure 3 and supplementary tables 1 and 2).

### Interrater reliability

Bland-Altmann plots showed no systematic differences in measurements of amplitude height between the two observers. The mean measurement difference (± SD) was 0.3 (± 2.2) µV for the lower amplitude and 0.9 (± 6.3) µV for the upper amplitude. The intraclass correlation coefficient was 0.83 for the upper amplitude and 0.87 for the lower amplitude.

## Discussion

This study is the first to provide evidence of varying aEEG amplitudes between wakefulness and sleep states in children. We observed the highest amplitudes during deep sleep. Amplitudes during light sleep were slightly lower than during wakefulness. With increasing age, amplitudes declined. The amplitude and bandwidth values give guidance to pediatric intensive care providers what upper and lower amplitudes and bandwidth to expect in unremarkable pediatric aEEGs.

The changes in amplitudes we observed can be explained by the raw EEG characteristics that define each sleep state: deep sleep (state N3) is characterized by high amplitudes and low frequencies (20). During wakefulness and sleep states N1 and REM, the EEG curve shows low voltages and high frequencies. Sleep state N2 is defined by high-amplitude K-complexes and sleep spindles (20). The voltage of the background activity can vary. This explains why sleep state 2 was detected during high and low amplitude episodes.

Episodes with high amplitudes were interrupted by amplitude dips. In critically ill comatose or sedated children, we have previously observed high amplitudes without the ability to interpret this finding (1). Some of these children displayed intermittent amplitude dips and some did not. Considering the results from this study, we assume that these dips can be interpreted as short transitions from deep to lighter sleep. Assessing aEEGs for amplitude dips during high-amplitude phases may help to identify patients with physiological sleep patterns in the PICU. Whether the lack of dips is associated with administration of sedation, specific clinical conditions or even outcomes in pediatric critical care patients, deserves further investigation.

A considerable proportion of patients in our study (40 %) received AEDs during the recording. The sensitivity analysis showed no differences in amplitude values and bandwidths between the entire cohort and patients without AEDs. This information is relevant for pediatric intensive care givers, because more than half of critically ill children undergoing continuous EEG monitoring in the PICU receive AEDs (23). Among those who later display seizures, 95 % are on AEDs at the time of EEG acquisition (23). We found no evidence in literature that sleep patterns are affected by AEDs, but children with epilepsy display sleep abnormalities more frequently than healthy children (24,25). Therefore, children admitted to the PICU due to seizure disorders may be a subgroup requiring special attention upon aEEG interpretation.

Our study has several limitations. The low case number, inclusion of patients with epilepsy, and wide age range prohibits to use the measured values as reference values for neurologically healthy children. Even though aEEG amplitudes did not differ between the whole cohort and patients without AEDs, the aEEG assessment may have missed differences in raw EEG activity between patients with and without AEDs. Patients receiving AEDs displayed higher percentages of sleep state N2 during high amplitude episodes compared to children without AEDs. In children with epilepsy, loss of physiological sleep patterns has been described, with higher proportions of N2 sleep (25). As this study excluded tracings with serious pathologies and was not designed to assess the sleep states during the entire tracing, the distribution of sleep states we observed in patients with AEDs is likely not representative for children with epilepsy. This makes further interpretation of our findings of sleep pattern distribution impossible.

This study is the first to provide evidence that aEEG amplitudes vary between wakefulness and different sleep states in stable children. Antiepileptic drugs did not seem to alter aEEG amplitudes. The prediction limits from this study provide intensive care givers with guidance what amplitude ranges to expect in awake and sleeping children: High amplitudes during deep sleep and lower amplitudes during wakefulness and light sleep. In children with epilepsy, the interpretation of aEEG amplitude variations requires special caution due to potentially disturbed sleep patterns.

## Data Availability

The datasets generated for this study will be made available upon request to any qualified researcher.

## Conflict of Interest

The authors declare that the research was conducted in the absence of any commercial or financial relationships that could be construed as a potential conflict of interest.

## Author Contributions

Study design: NB and ADM; conversion of aEEGs: VL, NB, and SG; analysis of aEEGs: VL and NB, raw EEG review for sleep state analysis: ADM; statistics: NB; drafting: NB; critical review and editing of the manuscript: UFM, CDS, HM, ADM, SG, VL.

## Funding

The study received funding from the Medical Faculty of the University of Duisburg-Essen (Corona Care-Program) and from the Stiftung Universitätsmedizin Essen. NB received an internal research grant from the Medical Faculty of the University of Duisburg-Essen (IFORES) and a grant from the Stiftung Universitätsmedizin Essen.

## Acknowledgments

N/A

---

**Supplementary Table 1:** Observed amplitude values for each channel of the sensitivity analysis (aEEGs conducted without antiepileptic drugs)

| Channel |  | Wakefulness |  |  | Sleep |  |  |  |  |  |
| --- | --- | --- | --- | --- | --- | --- | --- | --- | --- | --- |
| | | Lower ($\mu\text{V}$) | Upper ($\mu\text{V}$) | Bandwidth ($\mu\text{V}$) | High amplitude Lower ($\mu\text{V}$) | Upper ($\mu\text{V}$) | Bandwidth ($\mu\text{V}$) | Low amplitude Lower ($\mu\text{V}$) | Upper ($\mu\text{V}$) | Bandwidth ($\mu\text{V}$) |
| C3 – C4 |  | 20 (14 – 25) | 49 (36 – 59) | 29 (21 – 34) | 39 (27 – 44) | 101 (79 – 127) | 63 (50 – 84) | 17 (12 – 21) | 43 (32 – 52) | 26 (20 – 32) |
| P3 – P4 |  | 22 (17 – 25) | 51 (40 – 65) | 30 (23 – 39) | 38 (27 – 47) | 100 (74 – 124) | 53 (45 – 66) | 17 (14 – 20) | 44 (36 – 51) | 25 (22 – 31) |
| C3 – P3 |  | 16 (12 – 20) | 36 (29 – 48) | 21 (17 – 27) | 29 (20 – 36) | 87 (57 – 96) | 55 (37 – 61) | 14 (10 – 16) | 36 (27 – 46) | 22 (17 – 29) |
| C4 – P4 |  | 17 (13 – 21) | 43 (31 – 51) | 26 (17 – 29) | 30 (22 – 37) | 83 (59 – 102) | 53 (35 – 65) | 14 (11 – 16) | 39 (28 – 45) | 24 (18 – 29) |
| Fp1 – Fp2 |  | 19 (15 – 23) | 82 (64 – 94) | 60 (48 – 77) | 28 (22 – 32) | 83 (70 – 95) | 53 (45 – 66) | 10 (9 – 13) | 28 (25 – 33) | 17 (15 – 20) |

All values represent median and interquartile range

**Supplementary Table 2:** Sensitivity analysis: means and 95% confidence intervals for all patients vs. patients without antiepileptic drugs

|  |  | Wakefulness |  |  | Sleep |  |  |  |  |  |
| --- | --- | --- | --- | --- | --- | --- | --- | --- | --- | --- |
|  |  |  |  |  | High amplitude |  |  | Low amplitude |  |  |
| Channel |  | Lower (μV) | Upper (μV) | Bandwidth (μV) | Lower (μV) | Upper (μV) | Bandwidth (μV) | Lower (μV) | Upper (μV) | Bandwidth (μV) |
| C3 – C4 | All | 19 (17 – 21) | 47 (42 – 51) | 28 (25 – 31) | 36 (32 – 41) | 103 (90 – 116) | 67 (58 – 76) | 16 (15 – 18) | 41 (37 – 46) | 25 (22 – 28) |
|  | No AEDs | 20 (17 – 22) | 47 (41 – 53) | 28 (24 – 31) | 38 (32 – 43) | 104 (99 – 119) | 66 (57 – 76) | 17 (15 – 19) | 43 (37 – 49) | 26 (22 – 29) |
| P3 – P4 | All | 20 (18 – 22) | 51 (46 – 55) | 31 (28 – 33) | 36 (31 – 40) | 97 (85 – 108) | 61 (54 – 69) | 17 (15 – 18) | 44 (40 – 48) | 27 (25 – 30) |
|  | No AEDs | 21 (18 – 23) | 52 (45 – 58) | 31 (27 – 35) | 38 (32 – 43) | 100 (86 – 114) | 63 (54 – 71) | 17 (15 – 19) | 44 (40 – 49) | 27 (24 – 30) |
| C3 – P3 | All | 15 (13 – 17) | 37 (33 – 41) | 22 (19 – 24) | 28 (24 – 32) | 78 (68 – 88) | 50 (43 – 56) | 12 (11 – 14) | 34 (30 – 38) | 22 (19 – 25) |
|  | No AEDs | 16 (14 – 18) | 38 (32 – 43) | 22 (18 – 25) | 29 (25 – 33) | 79 (69 – 89) | 50 (44 – 56) | 13 (12 – 15) | 36 (32 – 41) | 23 (20 – 26) |
| C4 – P4 | All | 16 (14 – 18) | 40 (36 – 44) | 24 (21 – 26) | 29 (25 – 34) | 83 (71 – 94) | 53 (46 – 61) | 13 (12 – 15) | 37 (33 – 41) | 23 (21 – 26) |
|  | No AEDs | 17 (15 – 19) | 41 (35 – 46) | 24 (20 – 27) | 30 (25 – 34) | 82 (70 – 94) | 53 (45 – 60) | 13 (12 – 15) | 37 (33 – 41) | 24 (21 – 27) |
| Fp1 – | All | 17 (15 – 19) | 76 (67 – 86) | 58 (50 – 67) | 27 (24 – 30) | 82 (74 – 91) | 54 (49 – 61) | 11 (10 – 12) | 29 (25 – 33) | 18 (15 – 21) |
| Fp2 | No AEDs | 18 (16 – 20) | 81 (71 – 92) | 62 (53 – 72) | 27 (24 – 31) | 81 (72 – 92) | 53 (47 – 61) | 11 (10 – 13) | 30 (25 – 36) | 19 (15 – 23) |

Data represent mean and 95% confidence intervals

## Notes

### Competing Interest Statement

The authors have declared no competing interest.

